# Gestational Diabetes - Risk Factors and Outcomes among American Samoan Women (GROW): A Longitudinal Cohort Study Protocol

**DOI:** 10.1101/2025.10.24.25338761

**Authors:** Danielle J. Carson, Kima Faasalele-Savusa, Miracle Loia, Susie Tasele, Emele Iosefa, Peresia Tupuola, Kelly C. Sanchez, Oumaima Kaabi, Rachel K. Valencia, Mary G. Rossillo, Neharika Murthy, Tolulope Akinade, Lacey W. Heinsberg, Jenna C. Carlson, Clare A. Flannery, Stephen T. McGarvey, Ashlee N. Wood, Scott Anesi, Va’atausili Tofaeono, Katie Desobry, Bethel T. Muasau-Howard, Angela M. Bengtson, Erin E. Kershaw, Nicola L. Hawley

## Abstract

**Background:** Gestational diabetes mellitus (GDM) is linked with adverse health outcomes for both mother and infant and increases the risk of type 2 diabetes mellitus (T2DM) and long-term metabolic dysfunction postpartum. American Samoa experiences among the highest prevalence of GDM globally, with estimates suggesting 26-42% of women develop the condition. To date, no studies have attempted to understand the underlying etiology of GDM in Pacific Islanders or examine population-specific progression from GDM to T2DM. The GROW study will characterize glucose homeostasis, diabetes progression, and the impact of a Pacific-specific variant (rs373863828) in the *CREBRF* gene, which is known to protect against T2DM, on glucose metabolism during and after pregnancy.

**Methods:** We will establish a prospective cohort study of 350 pregnant women in American Samoa, enrolled in their first trimester and followed through 18 months postpartum. Participants will be genotyped for *CREBRF* rs373863828 and undergo frequently-sampled oral glucose tolerance test (fs-OGTTs), glycated hemoglobin (HbA1c), and continuous glucose monitoring (CGM) measurements to identify glycemic patterns across the perinatal period. Participants will complete questionnaires assessing reproductive health history, diet, physical activity, sleep, and psychosocial health. We will examine associations between *CREBRF* genotype and glucose homeostasis across pregnancy and postpartum and evaluate risk of GDM and subsequent T2DM. We will also explore associations between *CREBRF* genotype, changes in insulin secretion in pregnancy, and risk of adverse birth outcomes.

**Discussion:** Findings from this study are expected to inform precision medicine approaches to diabetes prevention, refine public health policies and clinical guidelines, and support community-based interventions aimed at reducing GDM and T2DM among Pacific Islander women and more broadly.

## Background

Gestational diabetes mellitus (GDM) is a well-established risk factor for future type 2 diabetes mellitus (T2DM) (1–3), with affected women facing up to a seven-fold higher risk of developing T2DM postpartum (4–6). GDM is linked to several pregnancy complications (7–9) and associated with lasting intergenerational effects, including macrosomia, excess adiposity, and long-term metabolic dysregulation in the child (10–12).

American Samoa, an unincorporated Pacific Island U.S. territory, experiences one of the highest burdens of GDM globally. Across the mainland U.S., GDM prevalence among Native Hawaiians and Pacific Islanders (NHPIs) was reported to be 10.6% (13) in 2020. In American Samoa, where >90% of residents identify as having Pacific Islander ethnicity, GDM prevalence is between 26-42% (13,14).Women in American Samoa comprise 49.2% of the total population (15) and are central in family life and community cohesion. However, cultural expectations to prioritize caregiving over personal well-being limit their time and ability to seek preventive healthcare, further compounding risk (16). Despite the elevated burden, no studies have attempted to understand the etiology of GDM, the population-specific risk of progression to T2DM associated with GDM, or to develop prevention and treatment interventions for this high-risk group.

Our previous research identified a population-specific novel missense variant, rs373863828 (p.Arg457Gln) in the *CREBRF* gene, with potential implications for GDM and T2DM prevention among Samoans and other Pacific Islanders (17). This missense variant, which is present among 45% of Samoans, is associated with greater obesity risk but paradoxically *lower* fasting glucose and reduced odds of T2DM (17), a finding that has been replicated across several other Pacific Islander populations (18–22). To date though, only one study has examined the *CREBRF* variant in the context of GDM (23). In 112 women of Māori and Pacific descent, there was a five-fold reduction in odds of GDM *per copy* of the A allele (OR: 0.19; 95% CI: 0.05, 0.69), independent of age, BMI, and family history. Although the study was limited in sample size and cross-sectional in nature, these findings suggest that the variant may have both a role in the etiology of GDM and utility as a marker of GDM risk. However, the mechanisms by which *CREBRF* impacts diabetes risk remain unknown.

Uncovering the mechanism by which *CREBRF* impacts diabetes risk has been challenging given the need for prospective studies and long latency period before diabetes onset. The rapid physiological changes that occur in pregnancy, however, including the natural emergence of insulin resistance in mid-to-late pregnancy (24,25), offer an exceptional “stress test” in which to examine the variant’s mechanism of action. One hypothesis that has not yet been adequately tested, is that the variant may be associated with enhanced first-phase insulin secretion (18); a finding that would potentially have broad pharmacological implications (26).

To address critical knowledge gaps about the mechanism of action of the *CREBRF* variant and its biomarker and the potential therapeutic target for diabetes risk we designed the Gestational Diabetes Risk Factors and Outcomes among American Samoan Women (GROW) study. This prospective, longitudinal study will follow women from early pregnancy (<14 weeks gestation) to 18 months postpartum, using gold-standard measures of glycemic control to explore pregnancy-related outcomes associated with the *CREBRF* variant, as well as its mechanisms of action and role in development of T2DM postpartum. Specifically, the study has four main aims: to determine (1) associations of *CREBRF* genotype with glucose homeostasis and GDM risk in pregnancy; (2) associations of *CREBRF* genotype with changes in glucose homeostasis and incident pre-DM/T2DM risk postpartum; (3) the extent to which improvements in insulin secretion mediate the pathway between *CREBRF* genotype and diabetes risk; and (4) to explore associations between genotype, changes in insulin secretion in pregnancy, and risk of adverse birth outcomes.

## Methods

### Study Design

GROW will enroll *n*=350 Samoan women in their first trimester of pregnancy and follow them through 18 months postpartum. Study visits will occur at 10-14 weeks’, 24-28 weeks’, and 32-38 weeks’ gestation and at 6 weeks, 6 months, 12 months, and 18 months postpartum. Participants will be genotyped for the *CREBRF* variant at their first study visit and will complete comprehensive assessments of glucose homeostasis, insulin secretion, body composition, and relevant covariates throughout the perinatal period. In addition, participants will complete a series of validated questionnaires capturing physical, mental, and behavioral health. Together, these measures will allow us to examine the interplay among metabolic, behavioral, and genetic factors, including the *CREBRF* variant, in determining GDM risk and progression to T2DM postpartum.

### Ethical Review

The GROW study received Institutional Review Board (IRB) approval from the Yale School of Public Health (Protocol #2000038646), Emory University, and the University of Pittsburgh with Yale serving as the IRB of record in a multi-site agreement. In American Samoa, the American Samoa Department of Health IRB granted their approval for the study as well as the Lyndon B Johnson (LBJ) Tropical Medical Center Research Oversight Committee.

### Study Setting

American Samoa is located approximately equidistant between Hawai’i and New Zealand. The majority of the population of ∼49,700 (27) is resident on the largest of several islands, Tuituila, which is home to the capital city of Pago Pago. Participants will be recruited from LBJ Tropical Medical Center, the only tertiary care hospital in the territory, and from four Federally Qualified Community Health Centers operated by the American Samoa Department of Health. The study will also be advertised on social media and promoted through collaboration with the local Women, Infants and Children (WIC) organization. Study visits and data collection will primarily take place at the Obesity, Lifestyle and Genetic Adaptations (OLaGA) Research Center, a partnership between Yale; the American Samoa Community College (ASCC); and the American Samoa Community Cancer Coalition (ASCCC), located in Nu‘uuli, American Samoa. Two of the postpartum visits (6-and 12-month assessments) may also take place in participant’s homes or in other private settings of their choosing.

### Study Population

Women will be eligible for the study if they are: (a) ≥18 years of age; (b) of Samoan ancestry (based on self-report of having four Samoan grandparents to ensure variation in the Pacific-specific variant rs373863828); (c) <14 weeks gestation based on ultrasound; have (d) a singleton pregnancy; (e) evidence of a completed prenatal care enrollment visit at either LBJ Hospital or one of the Federally Qualified Health Centers; (f) planning to reside in American Samoa until at least 18 months postpartum; and (g) willing and able to consent to research participation.

Exclusion criteria will include: (a) pre-existing Type 1 or T2DM, as defined by a self-reported diagnosis or fasting glucose ≥126 mg/dL or HbA1c ≥6.5% during the routine prenatal care enrollment visit, (b) conditions that would interfere with normal glucose homeostasis (e.g., medications, bariatric surgery), or with evaluation of study outcomes (e.g., congenital anomalies likely to affect fetal size), or key study variables (e.g., medical conditions interfering with adherence to continuous glucose monitoring (CGM) protocols, shift work that alters the sleep/wake periods and thus could likely affect the interpretation of CGM data), (c) use of glucose lowering medications or medications that significantly affect glucose homeostasis (i.e. steroids), or (d) skin allergies or conditions that would preclude the use of CGM monitors. History of GDM in prior pregnancies will be documented but will not be an exclusion criterion. Pre-diabetes (which we expect to be prevalent among the population) will also not be an exclusion criterion, as excluding this group would ignore a pressing public health need and limit generalizability.

### Recruitment and Retention

Recruitment will occur through clinic referrals, direct outreach (research assistants visiting prenatal care clinics), and social media, with eligibility confirmed after prenatal intake and provider referral. Eligible participants will complete an orientation/consent visit, during which bilingual staff will explain the study and obtain written informed consent. Retention strategies will include conducting home visits for assessments without significant biospecimen collection, providing appointment reminders and transportation, offering childcare during visits, maintaining engagement through social media updates, and offering incentives for study visits.

### Timeline

Participant recruitment began in April 2025 and is anticipated to be complete by September 2026. Data collection is anticipated to be completed by mid-2029. Interim data monitoring and preliminary analyses will be conducted on a rolling basis as study visits are completed. Final data cleaning and preparation for analysis are expected to occur by 2030, facilitating timely dissemination of study findings and manuscript preparation in the final phase of the project.

### Study Procedures

The GROW study includes three pregnancy and four postpartum visits. Visit-specific activities are summarized in **Table 1**. All study procedures will be conducted by trained, bilingual Samoan research staff.

**Table 1.**
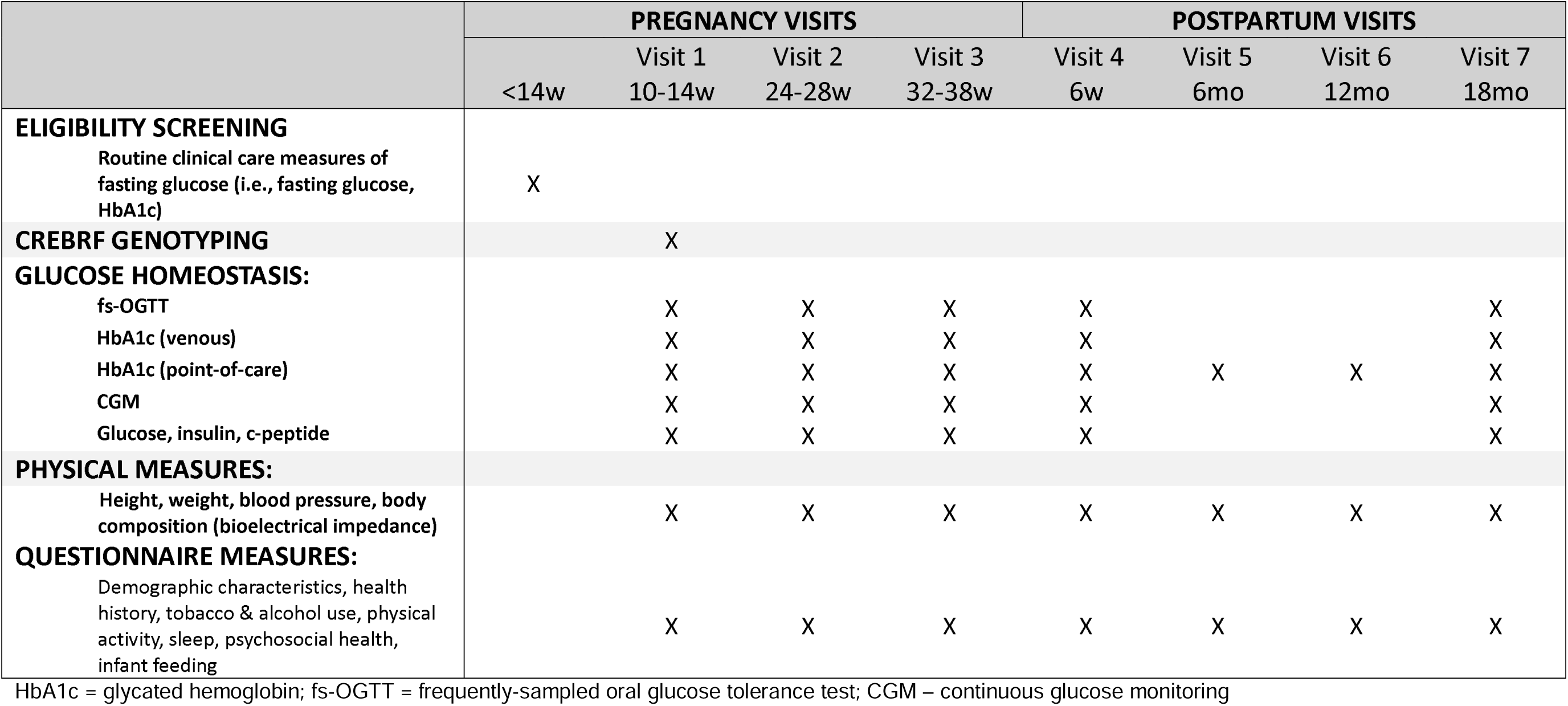
Overview of GROW Study Procedures.

### Glucose Homeostasis

We will comprehensively evaluate glucose homeostasis throughout the perinatal period using three complementary, well-validated approaches: (1) frequently sampled oral glucose tolerance tests (fs-OGTTs), (2) HbA1c, and (3) continuous glucose monitoring (CGM).

At each of the study visits indicated in **Table 1**, participants will complete a 2-hour, 75g fs-OGTT with blood sampling at baseline (pre-glucose) and six additional time points (15, 30, 45-, 60-, 90-, and 120-minutes post-glucose load). We will measure HbA1c in fasting samples and glucose, insulin, and C-peptide at each indicated time point, allowing us to calculate insulin secretion indices (28,29), insulin sensitivity (Matsuda Index) (30,31), and beta-cell adaptation through the Disposition Index (32,33). At 24-28 weeks’ gestation and 6 weeks’ postpartum, we will collect additional samples to be used for routine clinical GDM/T2DM screening, partnering with LBJ Hospital to avoid the need for participants to complete both routine and research assessments at these time points (a key retention strategy). We will also collect a point-of-care measure of HbA1c (A1cNow+, PTS Diagnostics, Whitestown, IN, USA) at all study visits, including at the 6- and 12-month postpartum visits, which will be used to provide participants with immediate feedback on their glycemic control.

Blood samples will be drawn from intravenous catheters into purple top vacutainer tubes containing potassium EDTA (whole blood for HbA1c), gray top vacutainer tubes containing sodium fluoride / potassium oxalate (plasma glucose), or red top vacutainer tubes containing silicone clot activator (serum insulin and C-peptide). The latter will be allowed to clot at room temperature for 30 min, whereas other tubes will be placed immediately on ice. Within 1 hour of collection, tubes will be centrifuged, and the resulting plasma or serum will be removed, aliquoted, and stored at −80°C until shipping on dry ice to the CLIA-certified University of Pittsburgh Medical Center (UPMC) Clinical Lab. Whole blood HbA1c levels will be determined using high performance liquid chromatography (HPLC) on a Tosoh G8 (Analytic Measurement Range (AMR) 4.3-17.0%; Percent Coefficient of Variation (%CV) ≤2.0). This method is certified by the National Glycohemoglobin Standardization program (NGSP) and standardized to the Diabetes Control and Complication Trial (DCCT assay). In addition, assessment of HbA1c from frozen whole blood has been validated (34,35). Serum insulin (AMR 0.4111000 μU/mL or 2.78116945 pmol/L; %CV ≤2.0) and C-peptide (AMR 0.0071113.3 nmol/L or 0.021140 ng/mL; %CV ≤3.2) will be determined using electrochemiluminescence Immunoassays (ECLIA, Elecsys Reagent Packs) on a Roche Cobas e801. Plasma glucose will be determined by an enzymatic hexokinase method in a Beckman Coulter AU5800.

Continuous glucose monitoring (CGM) will be conducted as part of the five study visits shown in **Table 1**. Freestyle Libre Pro devices (Abbot Laboratories, Lake County, Illinois) will be placed on the back of the upper arm by research staff the day before participants attend research assessments. The sensor, which records a measurement of glucose levels in the participant’s interstitial fluid every 15 minutes, will be monitored for the first hour of wear using the reader device to ensure correct placement, after which participants will be instructed to continue with their usual activities until the device is removed by study staff 10 days after the research assessment (∼11 days total wear time). Participants will be blinded to the CGM data. CGM data will be analyzed using functional data analysis to generate standard glycemic metrics and 24-hour glucose profiles. CGM is FDA approved for use in pregnancy and was selected for its ability to provide low-burden and real-time data that can reveal glycemic dysregulation that may not be captured by HbA1c or the fs-OGTT (36–39).

### *CREBRF* Genotyping

During the baseline draw of the 10–14-week fs-OGTT, whole blood will be collected in a 2.5 mL in Vitro Diagnostics (IVD) PAXgene^®^ Blood DNA tube (PreAnalytiX GmbH, catalog #761165). After thorough mixing, whole blood will be aliquoted, then stored at −80°C before transportation to the UPMC for DNA extraction using QIAamp DSP DNA Blood Mini Kit (PreAnalytiX GmbH, catalog #61104). Resulting DNA will then be genotyped for the CREBRF rs373863828 SNP using a validated pre-designed TaqMan^®^ SNP assay (Applied Biosystems; Assay ID# C_203097374_10, catalog #4351379) on an ABI 7900HT instrument (40). TaqMan™ assay (Applied Biosystems, Waltham, Massachusetts). Empirical kinship and ancestry among the sample (to control for relatedness) will be evaluated using the iPLEX^®^ Pro Sample ID Panel on a MassARRAY^®^ System (Agena Biosciences, # 13116F) in the UPMC Clinical Genomics Center.

### Physical Measures

At each study visit, height and weight will be measured in duplicate using a portable stadiometer (SECA, Hamburg, Germany) and digital scale (Tanita Corporation of America, IL, USA), respectively. Measures will be used to calculate body mass index (BMI). Body composition will be assessed using segmental multi-frequency bioelectrical impedance (Seca mBCA 554, Hamburg, Germany), which estimates muscle mass, fat mass, and body water for each limb and the trunk region. Measurements will follow pregnancy-specific guidelines to maximize longitudinal validity by collecting all measures after a standard fasting period, at a similar time of day, and after asking participants to empty their bladder (41). Blood pressure will be measured in triplicate after a 5-minute seated rest, with 3-minute rest periods between readings, using an Omron HEM-907XL monitor (Omron Healthcare, Muko, Kyoto, Japan).

### Questionnaire Measures

At each study visit, participants will complete questionnaires that will collect data on demographic characteristics, health history, tobacco and alcohol use, dietary intake, physical activity, sleep, household characteristics, and psychosocial health. Postpartum visits will also collect information about infant feeding practices, child development, and parenting. Questionnaire Measures are summarized in **Table 2**. Measures related to medical conditions and interventions, income, financial assistance, and strain, and stressful life events during pregnancy were adapted from the Environmental Influences on Child Health Outcomes (ECHO) study (42). Each of the questionnaires has been translated into the Samoan language. Bilingual research assistants will administer questionnaires and record responses in REDCap (43–45).

**Table 2.**
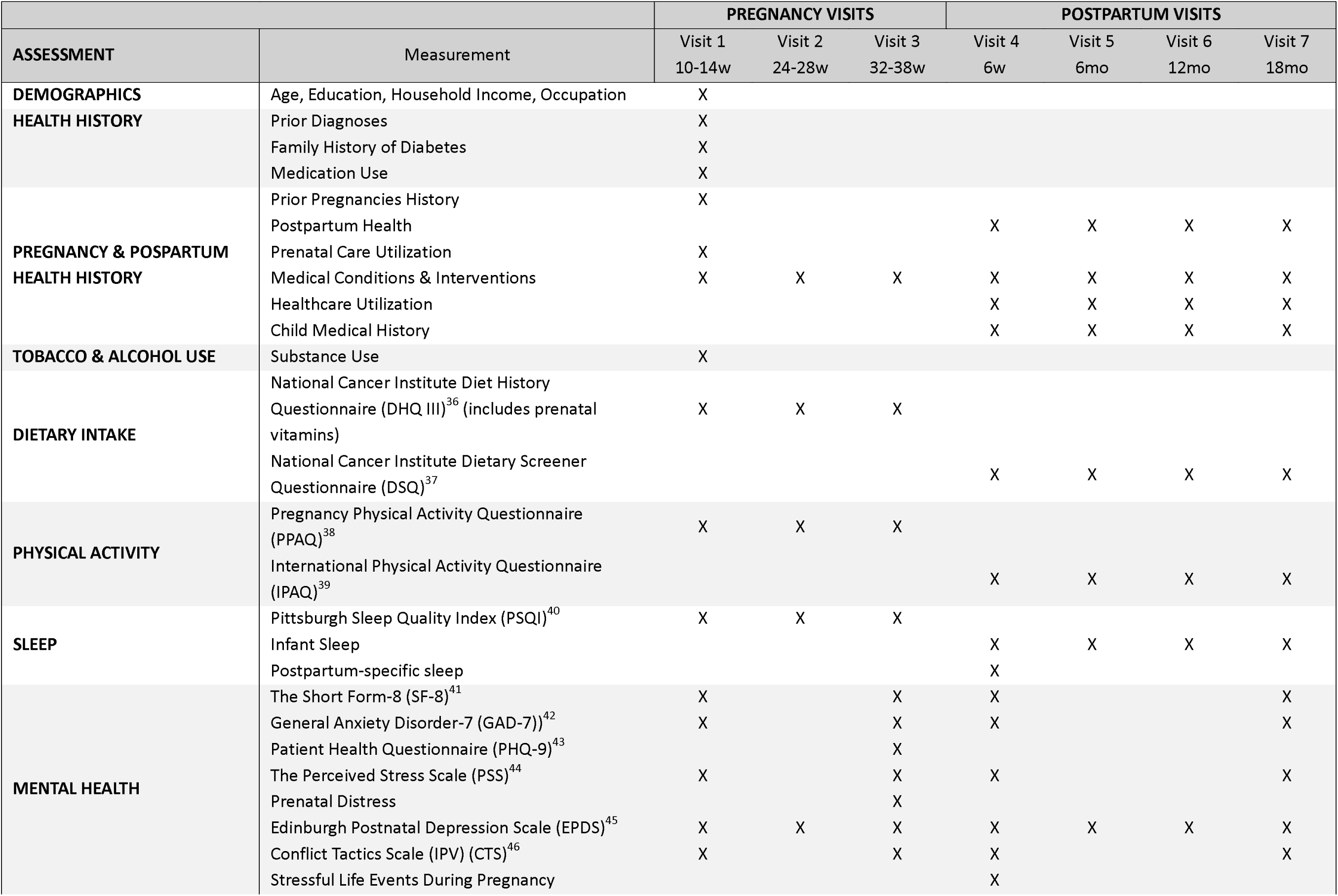
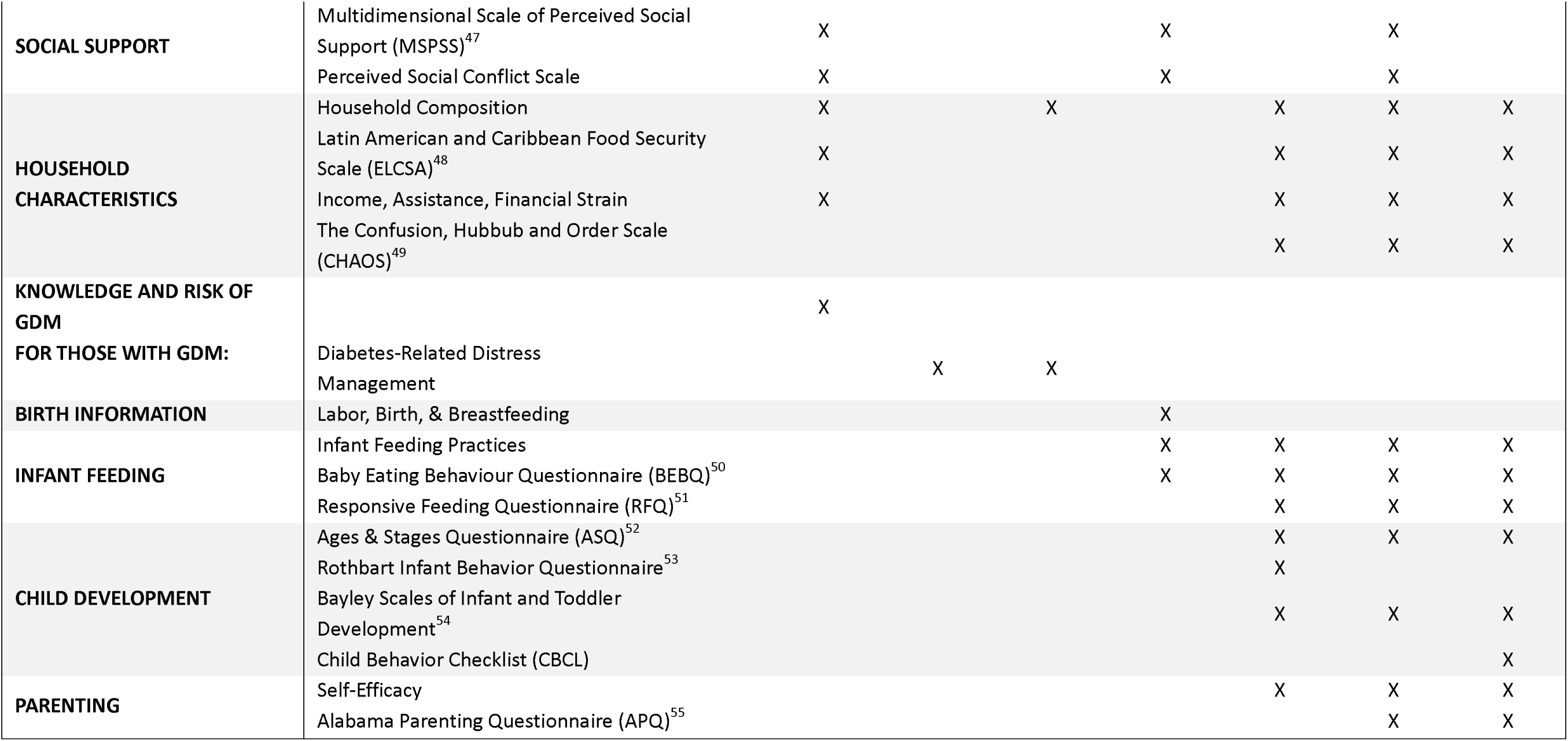
GROW Questionnaire Measures.

### Biobanking

We will ask participants permission to biobank any remaining biospecimens from each of the blood draws completed as part of the study. Participants will have the option to decline biobanking and still be included in the study. Banked biospecimens will include whole blood, serum, plasma, and DNA. Biospecimens will be stored at −80°C at the University of Pittsburgh for use in future research.

### Medical Record Review

Clinical data will be extracted from participants’ medical records to confirm eligibility, supplement questionnaire responses, and provide information about GDM diagnosis. We will also extract data on other pregnancy diagnoses (such as preeclampsia), GDM treatment, and infant outcomes, including birth weight, gestational age, delivery mode, and neonatal hypoglycemia diagnosis.

### Participant Feedback & Referrals

At each assessment participants will receive information about their physical measurements, blood pressure, HbA1c, and dietary intake. We will remind participants that all information collected is for research purposes and does not replace clinical care. Health system referrals will be provided for any participant who exhibits high blood pressure (>140/90 mmHg for two or more readings), low blood pressure (<90/60 mmHg), HbA1c ≥6.5%, or an Edinburgh Postnatal Depression Scale (EPDS) score ≥13. Any participant endorsing thoughts of self-harm (a positive response on EPDS item 10) or suicidal ideation will be assessed and referred to mental health services. Participants may request information about their *CREBRF* genotype, although we will be careful to explain that our research quality genotyping is not equivalent to a CLIA-certified assessment. We will also explain to participants that the information that we know about the *CREBRF* variant is not currently actionable and that advice to maintain a healthy lifestyle and seek regular screening for diabetes, would be the same regardless of genotype.

### Data Management & Quality

Data will be collected by trained OLaGA research center staff and stored in a secure REDCap database on a Yale University cloud server, with daily de-identified uploads to collaborating institutions. Dedicated data managers will implement quality control procedures throughout the study, with regular assessment of missing values, inconsistent records, and outliers. Research staff will receive standardized training with ongoing re-training and reliability checks for physical measurements. Laboratory analyses will be conducted at UPMC’s clinical lab to ensure standardization and adherence to clinical guidelines.

### Data Sharing

During the informed consent process, participants will have the option to opt in or out of broad sharing of study data and biospecimens, including public data sharing, in accordance with NIH policy. Consent preferences will be recorded in REDCap to ensure that only data from participants who authorize broad sharing are submitted. For those who consent, de-identified data will be deposited in the Database of Genotypes and Phenotypes (dbGaP) federal registry under a unique accession number. De-identification will follow Health Insurance Portability and Accountability Act (HIPAA) privacy standards, and data will be maintained indefinitely to support transparency and respond to future inquiries related to published findings. To access the data, researchers must follow the established dbGaP request process, subject to approval by the dbGaP access committee. Given the inclusion of individuals from a small and geographically isolated population, we will request additional access limitations to protect participant privacy. Specifically, we will require that requestors: (1) provide documentation of IRB approval from their institution and the American Samoan IRB, (2) commit to sharing results with the broader scientific community, (3) submit a letter of collaboration with the study’s principal investigators, and (4) be affiliated with not-for-profit institutions.

To further protect participant privacy, biospecimens will be labeled only with a unique study ID and date or visit number. Biospecimens will be securely stored at the University of Pittsburgh in alarm-monitored freezers within a restricted-access laboratory. Only authorized study personnel who have completed appropriate training will have access. These protections, along with institutional oversight by Yale, Emory, and the University of Pittsburgh, ensure rigorous stewardship of data and uphold participant confidentiality throughout the data sharing process.

### Statistical Analysis Plan

Primary study outcomes include GDM at 24-28 weeks, changes in HbA1c and fasting plasma glucose between 10-14 weeks gestation and 18 months postpartum, incident pre-diabetes and T2DM at 18 months postpartum, and assessments of both insulin secretion and insulin action across pregnancy and postpartum. Secondary study outcomes will examine the extent to which first-phase insulin response mediates associations between *CREBRF* genotype and changes in HbA1c, fasting plasma glucose, and GDM and pre-DM/T2DM risk. We will also explore the extent to which improved insulin secretion (anticipated among those with the *CREBRF* variant) may adversely affect birth outcomes (abstracted from the medical record), specifically large-for-gestational age (based on birth weight), birth by cesarean section, and neonatal hypoglycemia.

We will primarily use an additive genetic model (GG=0, AG=1, AA=2) to assess how the *CREBRF* variant influences glucose homeostasis, GDM risk, and T2DM progression in women, and adverse birth outcomes in infants. Descriptive analyses will summarize categorical and continuous variables using appropriate measures. Multivariable logistic or linear regression will estimate associations between genotype group and GDM/T2DM risk and markers of glucose homeostasis (fasting glucose, HbA1c, etc.) during the perinatal and postpartum period (46). CGM data will be used to characterize glucose patterns over time to identify differences by genotype and identify clinically relevant glucose and insulin dynamics (39). Causal mediation analyses will be used to evaluate the extent to which improved insulin secretion and body composition mediate associations between genotype with GDM/T2DM diabetes risk and glucose homeostasis (47). Finally, logistic or linear regression and mediation analyses will be used to explore associations between maternal *CRBERF* genotype, maternal insulin secretion during pregnancy, and adverse birth outcomes (large-for-gestational-age [LGA], cesarean delivery, and neonatal hypoglycemia). Models will be adjusted for a list of covariates identified *a priori* including baseline weight, body composition, age, family history of diabetes, parity, previous LGA birth, diet, physical activity, socioeconomic status, and comorbidities as well as gestational weight gain and postpartum changes in body composition where applicable. Missing data will be handled with multiple imputation if necessary, and inverse probability censoring weights will adjust for pregnancy loss and participant loss to follow up. Sensitivity analyses will assess alternative model specifications, impact of missing data, and potential unmeasured confounding. The study is powered at over 90% to detect meaningful differences in outcomes. For primary analyses, we will use 2-sided tests at the 5% significance level. Statistical analyses will be performed using R or Stata.

## Discussion

This study will be among the first prospective cohort studies to examine the etiology of GDM and the transition from GDM to postpartum T2DM in Pacific Islanders, with a central focus on the *CREBRF* genetic variant. *CREBRF* rs373863828 is uniquely prevalent in Pacific Islanders and has been associated with increased adiposity, enhanced insulin secretion, and lower fasting glucose, yet its impact on glucose homeostasis during pregnancy and postpartum remains unclear. By integrating CGM and fs-OGTTs, this study will generate a comprehensive metabolic profile across the perinatal period, helping determine the extent to which *CREBRF* rs373863828 can serve as a genetic biomarker for diabetes risk and guide targeted prevention strategies for high-risk populations. In addition, by generating the first ever population-specific longitudinal CGM data during pregnancy and postpartum, this study will contribute to ongoing research exploring CGM’s role in diabetes prediction (48). Finally, we anticipate that longitudinal analytical methods, paired with these unique data, will allow us to uncover novel genotype-specific associations, providing new insights into diabetes progression and prevention.

Our expectation is that findings from this work will advance precision medicine approaches to diabetes prevention and improve long-term health outcomes for the high-risk Samoan population. Findings from this study will inform an improved biological understanding of *CREBRF* that may inform public health policies, refine clinical practice guidelines, and support community-based interventions in American Samoa, ultimately working toward reducing diabetes disparities and improving maternal and infant health outcomes in Pacific Islander communities and beyond. Although the *CREBRF* variant is predominantly found in Pacific Islanders, results from this study will also help clarify mechanisms underlying obesity-related metabolic disease risk in other high-burden populations.

## Data Availability

This protocol presents no data.

## Acknowledgements

The authors would like to express their appreciation to the American Samoa Department of Health leadership for the support of their study. We would also like to thank the clinical and administrative staff at the Lyndon B Johnson Medical Center, particularly Dr. Efren Yuchongco, and the DOH clinics (Drs. Mirella, Adriano, and Sema) for their support in recruitment. Finally, we acknowledge the logistical support provided by staff at the American Samoa Community Cancer Coalition (Tulimalefoi Vaofanua) and American Samoa Community College (Mark Schmaedick).

## Declarations

### Availability of data and materials

Participants will have the option to consent to broad data sharing through dbGaP. The dbGaP accession number will be reported in all study manuscripts.

### Author Contributions

Danielle Carson – Writing – original draft, Writing – review & editing; Kima Faasalele-Savusa – Methodology, Project administration, Supervision, Writing – review & editing; Miracle Loia – Project administration, Writing – review & editing; Susie Tasele - Project administration, Writing – review & editing; Emele Iosefa - Project administration, Writing – review & editing; Peresia Tupuola - Project administration, Writing – review & editing; Kelly Sanchez – Methodology, Project administration, Supervision, Writing – review & editing; Oumaima Kaabi – Methodology, Project administration, Writing – review & editing; Rachel Valencia – Methodology, Project administration, Writing – review & editing; Mary Rossillo – Methodology, Project administration, Writing – review & editing; Neharika Murthy – Writing – review & editing; Tolulope Akinade –Project administration, Writing – review & editing; Lacey Heinsberg – Conceptualization, Funding acquisition, Methodology, Project Administration, Writing – review & editing; Jenna Carlson - Conceptualization, Funding acquisition, Methodology, Project Administration, Supervision, Writing – review & editing; Clare Flannery - Conceptualization, Funding acquisition, Methodology, Project Administration, Writing – review & editing; Stephen McGarvey - Conceptualization, Funding acquisition, Methodology, Project Administration, Writing – review & editing; Ashlee Wood - Methodology, Project Administration, Writing – review & editing; Scott Anesi – Resources, Writing – review & editing; Va’atausili Tofaeono – Resources, Methodology, Writing – review & editing; Katie Desobry – Methodology, Writing – review & editing; Bethel Muasau-Howard - Conceptualization, Funding acquisition, Methodology, Project Administration, Supervision, Writing – review & editing; Angela Bengtson - Conceptualization, Funding acquisition, Methodology, Project Administration, Supervision, Writing – original draft, Writing – review & editing; Erin Kershaw - Conceptualization, Funding acquisition, Methodology, Project Administration, Supervision, Writing – original draft, Writing – review & editing; Nicola Hawley - Conceptualization, Funding acquisition, Methodology, Project Administration, Supervision, Writing – original draft, Writing – review & editing.

